# Can Language Models Diagnose Sepsis? A Head-to-Head Evaluation Against 353 Physicians

**DOI:** 10.64898/2026.10.23.26366291

**Authors:** Eric Macias-Fassio, David Betancur, Cristina Pruenza, Andres Giglio, Giuliano Gallano, Marcio Borges-Sa, Aythami Morales, Julian Fierrez

**Affiliations:** Instituto de Ingeniería del Conocimiento, Madrid, Spain; Multidisciplinary Sepsis Group, Son Llatzer Hospital Palma, Palma, Spain; Health Research Institute of the Balearic Islands (IdISBa), Palma, Spain; Balearic Islands University, Palma, Spain; Fundación Código Sepsis, Spain; Faculty of Medicine, Finis Terrae University, Santiago, Chile; Critical Care Center, Clínica Las Condes Hospital, Santiago, Chile; Departamento de Matematicas, Universidad de las Palmas de Gran Canaria, Spain; BiometricsAI, Universidad Autónoma de Madrid, Madrid, Spain

**Author notes:** **Email address:** (Eric Macias-Fassio).

**Keywords:** Sepsis, Large Language Models (LLMs), Decision Support System

## Abstract

**Importance:** Sepsis is a leading cause of in-hospital mortality, where timely and accurate diagnosis is critical yet frequently delayed. Whether large language models (LLMs) can diagnose sepsis reliably — and whether they do so fairly, consistently, and with well-calibrated confidence — remains unknown.

**Objective:** To evaluate the diagnostic capabilities of LLMs on standardised sepsis vignettes before and after biomarker disclosure, benchmarked against physicians, and to characterise their robustness, stability, bias and confidence calibration.

**Design:** Retrospective in-silico study based on six anonymised real cases spanning the diagnostic spectrum from no infection to septic shock. For each case, the reference-standard diagnosis was assigned, following the Sepsis-3 framework. Each case was evaluated under a two-stage (pre– and post–biomarker disclosure) protocol. To assess potential demographic bias, every case was additionally presented while systematically varying the patient’s sex, age, and language.

**Setting and Participants:** Five locally deployed LLMs (DeepSeek-4, Qwen3.8-27B, its quantised variant, Gemma-4-31B, and the domain-specific MedGemma) were evaluated in a controlled evaluation pipeline and compared with responses from 353 clinicians, using identical clinical vignettes.

**Main Outcomes and Measures:** Diagnostic accuracy before/after biomarker disclosure, per-vignette error, sensitivity to demographic attributes (diagnosis-change rate) and its directional component, reproducibility across repetitions and confidence calibration.

**Results:** Before biomarkers, model accuracy ranged 40–67%, comparable to physicians (57%). After disclosure, physician accuracy was 61%, exceeded by Qwen3.8-27B (91%), its quantised variant (86%) and Gemma-4-31B (83%); DeepSeek-4 reached 64% and MedGemma 56%, the only model below physicians. Biomarkers rescued confirmatory cases (pancreatitis-associated sepsis), whereas post-surgical sepsis remained difficult for models and physicians alike. Quantisation cost only 5 accuracy points. Patient sex and age showed no directional bias, and language was the only bias. Gemma-4-31B was the most reproducible model, the Qwen models were the most accurate and best calibrated, and MedGemma the least.

**Conclusions and Relevance:** LLMs, including quantised variants, can provide accurate informed decision support for sepsis, surpassing physicians once laboratory data are available. However, robustness, calibration, and language fairness varied independently of accuracy and should guide model selection and clinical validation.

## 1. Introduction

Sepsis is a life-threatening syndrome characterized by organ dysfunction resulting from a dysregulated host response to infection, including bacterial, viral, and fungal infections [1]. It represents a major global health burden, with an estimated 48.9 million cases and 11 million sepsis-related deaths occurring worldwide each year [2]. Because sepsis is a time-sensitive syndrome, early recognition and treatment are essential for improving patient outcomes. Several studies have shown that machine-learning (ML) models can facilitate earlier identification of patients at risk of sepsis and may improve clinical outcomes [3, 4]. However, models deployed in clinical settings that differ from those in which they were developed often exhibit limited generalizability [5, 6]. A major source of heterogeneity is the lack of a universally applied operational definition of sepsis. Hospitals and healthcare systems may use Sepsis-2 or Sepsis-3 criteria, clinical scoring systems, expert judgment, or combinations of these approaches [7]. This variability complicates both clinical diagnosis and the development and external validation of predictive models. Consistent with this challenge, previous studies have reported limited diagnostic accuracy and substantial disagreement among clinicians when identifying and classifying sepsis [8, 9, 10].

To address this issue, our previous study evaluated six anonymized clinical cases in a multinational cohort of clinicians [11]. We assessed the accuracy of their diagnoses against a predefined reference standard and examined the effect of providing additional biomarker information. Building on these findings, the present study systematically evaluates the diagnostic performance of several state-of-the-art large language models (LLMs) using the same clinical cases and survey framework, thereby enabling a direct comparison between LLMs and clinician diagnoses. LLMs are computational models trained on extensive text corpora to learn complex linguistic patterns, enabling them to perform a wide range of tasks beyond those explicitly represented during training. An important concern is that LLMs may be trained on data that include human biases, which could alter their performance in less-represented subgroups. Some simulation studies and meta-analyses have found that biases may arise in relation to gender or ethnicity, leading LLM performance to vary or deteriorate across certain subgroups. This may disadvantage specific populations and contribute to health disparities [12, 13, 14].

In the present study, we analyze the diagnostic performance of LLMs in challenging sepsis cases and compare it with that of a multinational cohort of clinicians (N=353). We also conduct an extensive analysis of potential LLMs biases by examining whether diagnostic performance varies according to patient age, sex, and the language in which the clinical cases are presented.

## 2. Methodology

### 2.1. Clinical Cases

Six vignettes were developed by three senior clinicians from the Multidisciplinary Sepsis Unit at Hospital Universitario Son Llàtzer, based on real, fully anonymised patients with documented clinical course and outcome. For each case, only information available within the first 24 hours of presentation was included in the vignette text shown to respondents. The case set was designed to cover the diagnostic spectrum of suspected infection in critically or seriously ill adults. It included one negative control vignette, representing infection without sepsis; one positive control vignette, representing septic shock; and four diagnostically challenging vignettes involving non-infectious confounders, immunocompromise without confirmed infection, and sepsis related to surgical or urological pathology (Appendix A). A reference diagnosis was assigned to each vignette by a panel of four clinicians from intensive care, internal medicine, and infectious diseases, including one clinician external to the participating centres, based on the patient’s documented clinical course, including subsequent clinical evolution, laboratory trajectory, and microbiological findings, applying the Sepsis-3 framework [1].

### 2.2. Survey structure

The survey first captures the sepsis definition used by the clinicians and the LLMs (Sepsis-2, Sepsis-3, combination of definitions, expert judgment or unsure). For each case, LLMs and clinicians first received the clinical history, examination findings, basic laboratory results, and imaging available within the first 24 hours, and were asked to (i) select a diagnosis from five options (no infection, infection without sepsis, sepsis, severe sepsis, or septic shock), (ii) rate their diagnostic certainty on a five-point ordinal scale (very low, low, moderate, high, very high), and (iii) optionally provide a free-text justification. Biomarker results were then disclosed; the biomarkers shown were selected to reflect those typically available in the clinical scenario portrayed and included procalcitonin, C-reactive protein, white blood cell count and differential, and lactate, with monocyte distribution width additionally provided where institutional availability of this parameter was clinically realistic. Following biomarker disclosure, LLMS and clinicians were asked to (iv) provide a final diagnosis, (v) rate their final certainty on the same scale, and (vi) indicate whether the biomarker information had been useful for their diagnostic decision. The response option “severe sepsis” was retained to accommodate clinicians and LLMs who use Sepsis-2 terminology and was merged with “sepsis” for analytic purposes, consistent with the Sepsis-3 framework. The survey is reported in accordance with the CHERRIES checklist [15].

### 2.3. Large Language Models

We evaluated five LLMs, all deployed locally. The panel was chosen to span dense and Mixture-of-Experts (MoE) architectures, different model scales and generations, general-purpose versus finetuned on medical domain, and full-precision versus quantised weights.

#### *Gemma-4-31B.* (google/gemma-4-31B-it [16])

This is the flagship dense model of Google DeepMind’s Gemma 4 family [17], with 30.7B parameters and a 256K token context window, released in April 2026 under an Apache 2.0 license with reasoning and multimodal capabilities. We used the instruction-tuned checkpoint. It was included as a current-generation, general-purpose dense model of similar scale to MedGemma.

#### *DeepSeek-V4.* (deepseek-ai/DeepSeek-V4-Flash-0731 [18])

This is the official release of DeepSeek-V4-Flash, published on 31 July 2026 with MIT-licensed weights [19]. It is a sparse MoE model with 284B total parameters, 13B active per token and a one-million-token context window. Like the rest of the V4 family, it integrates reasoning into a single model with switchable thinking and non-thinking modes. It is the largest model in our panel by total parameter count.

#### *MedGemma.* (google/medgemma-27b-text-it [20])

This is Google’s domain-specific variant of Gemma for medical applications [21], released in May 2025 and built on the Gemma 3 backbone [22]. It is the only medically specialised model in the panel and therefore tests whether domain adaptation confers an advantage in sepsis diagnosis over newer general-purpose models.

#### *Qwen3.8-27B.* (Qwen/Qwen3.8-27B [23])

This is Alibaba’s 27B dense open-weight model, released on 14 August 2026 under Apache 2.0. It is a native vision–language model with adjustable reasoning, positioned by its developers for single-GPU and on-premise use. It is the most recent model evaluated.

#### *Qwen3.8-27B (4-bit).* (Qwen/Qwen3.8-27B [23], 4-bit)

This is a 4-bit weight-quantised version of the same Qwen3.8-27B checkpoint. Rather than using a separate pre-quantised release, the original BF16 weights were quantised in-flight at load time with bitsandbytes [24], as supported by vLLM [25]. This method requires no calibration data, so the quantised model differs from the full-precision one only in the numerical precision of its weights. Quantisation reduces weight memory to roughly a quarter of the BF16 model, allowing it to run on a smaller GPU. It was included to isolate the effect of quantisation on accuracy, robustness and bias while holding architecture and training constant, reflecting the deployment scenario most realistic for resource-constrained clinical settings.

### 2.4. Bias Analysis

To assess potential sources of bias in the LLMs, each clinical case was evaluated in both Spanish and English. For each case, simulations were conducted for male and female patients across seven age groups (25, 35, 45, 55, 65, 75, and 85 years). Furthermore, evaluations were performed with the reasoning (“thinking”) mode both enabled and disabled, and at three temperature settings (0.0, 0.3, and 0.7). To assess response stability, each experimental configuration was repeated three times. Overall, the study design comprised 6 clinical cases x 2 languages (Spanish and English) × 7 age groups × 2 genders × 2 reasoning modes × 3 temperature settings × 3 repetitions, resulting in 3,024 inference scenarios. Because each scenario was evaluated at two stages (before and after biomarker disclosure), a total of 6,048 LLM calls were performed.

### 2.5. Statistical Analysis

Diagnostic accuracy was computed as the proportion of correct diagnoses against the Sepsis-3 reference standard, before and after biomarker disclosure. Proportions are reported with 95% Wilson confidence intervals. For each demographic factor (patient sex, age and language), sensitivity was quantified as the diagnosis-change rate (the proportion of otherwise-identical configuration groups in which the voted diagnosis changed when only that factor varied). To distinguish directional bias from non-directional noise, we contrasted this rate with the accuracy spread across the factor’s levels (maximum *−* minimum accuracy). Within-model reproducibility was defined as the proportion of configurations in which all three repetitions produced an identical diagnosis, computed overall and stratified by sampling temperature. Calibration was assessed by computing accuracy within each self-reported confidence level.

## 3. Results

### 3.1. Diagnostic Accuracy

Figure 1 reports diagnostic accuracy for the five LLMs alongside the physician baseline. Before biomarker disclosure, model accuracy pooled across all configurations ranged from 40% to 67%, broadly comparable to physicians (57%). Disclosure changed this picture markedly: the three strongest models surpassed the physician comparator, with Qwen3.8-27B reaching 91% (+26 points), its quantised variant 86% (+22), and Gemma-4-31B 83% (+16). DeepSeek-4 and MedGemma improved more modestly, to 64% (+19) and 56% (+16) respectively, with MedGemma remaining the only model below the physician baseline. Notably, quantisation cost Qwen3.8-27B only five accuracy points (91% → 86%), indicating that the quantised model retained most of its diagnostic performance.

**Figure 1:**
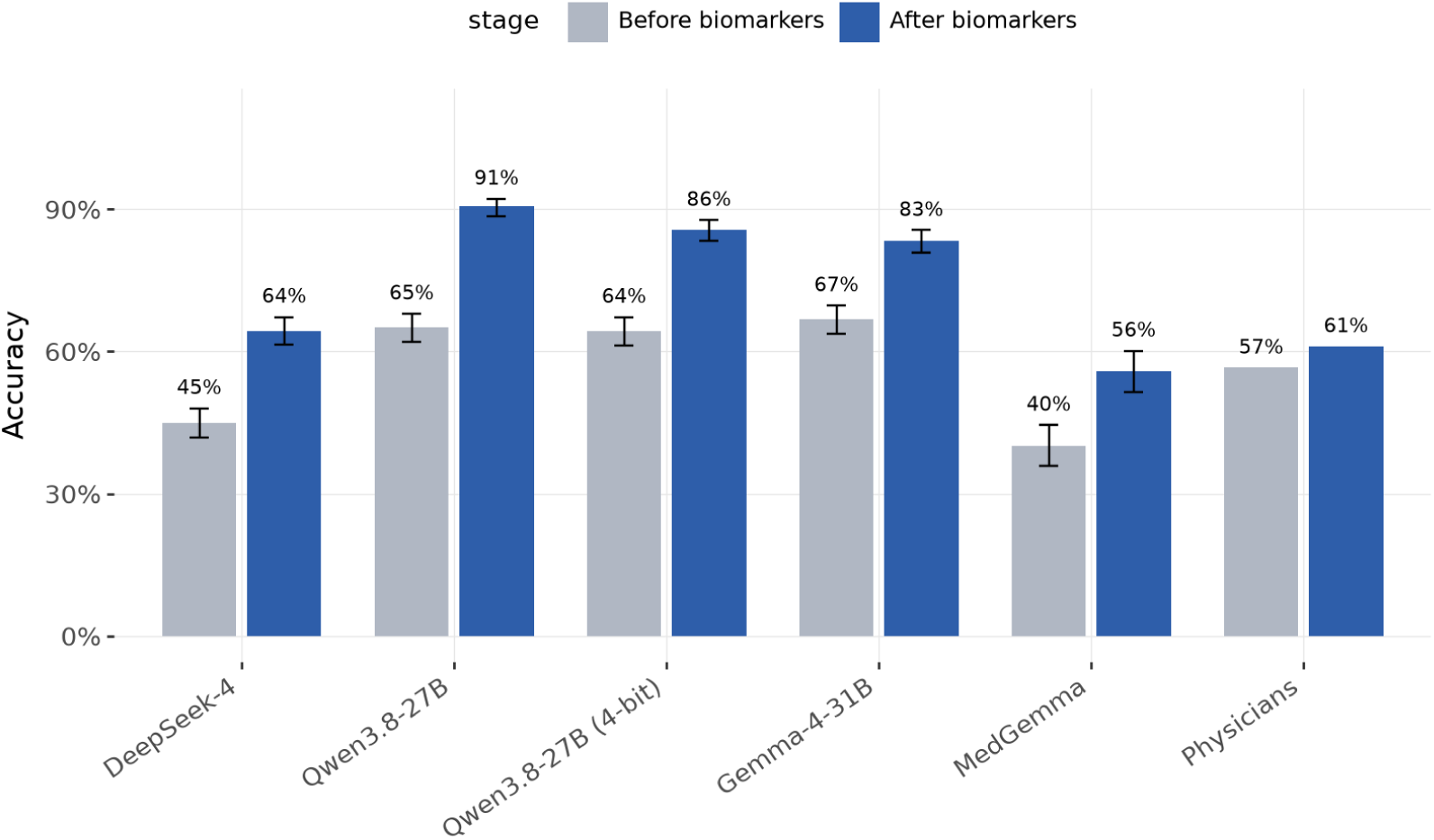
Diagnostic accuracy of each LLM before and after biomarker disclosure, compared with physicians. Bars show the proportion of correct diagnoses (against the Sepsis-3 reference standard) for each model, pooled across all experimental configurations — patient sex, age, language, reasoning mode and sampling temperature, with the three repetitions of each configuration collapsed by majority vote (N = 1,008 configurations per model; MedGemma N = 504, as it has no reasoning-mode toggle). The physician comparator (N = 353 physicians) is the mean accuracy across the six clinical vignettes of the companion survey [11] and serves as the baseline reference.

A per-vignette breakdown (Figure 2) clarifies where these gains originated. The two control cases — Case 1 (infection without sepsis) and Case 2 (septic shock) — were solved by all models except MedGemma, with 0% error in both stages. The diagnostically challenging sepsis cases, by contrast, were frequently missed before biomarkers: the larger models erred in 98–100% of configurations on Case 3 (sepsis in acute pancreatitis) and in 96–100% on Case 5 (post-surgical sepsis), reflecting a systematic tendency to under-recognise sepsis when confirmatory laboratory data were withheld. Biomarker disclosure then produced a striking, case-specific rescue: on Case 3 the error rate collapsed from 98–100% to 0–14%, showing that the information needed to reclassify the case was carried almost entirely by the biomarkers. Case 5, however, remained the hardest vignette even after disclosure, with most models still misclassifying it; physicians were likewise poor here (80% → 64%), suggesting that post-surgical sepsis is intrinsically difficult rather than a model-specific failure. MedGemma was the exception, showing an anomalous error profile that did not follow the clinical difficulty gradient of the other models.

**Figure 2:**
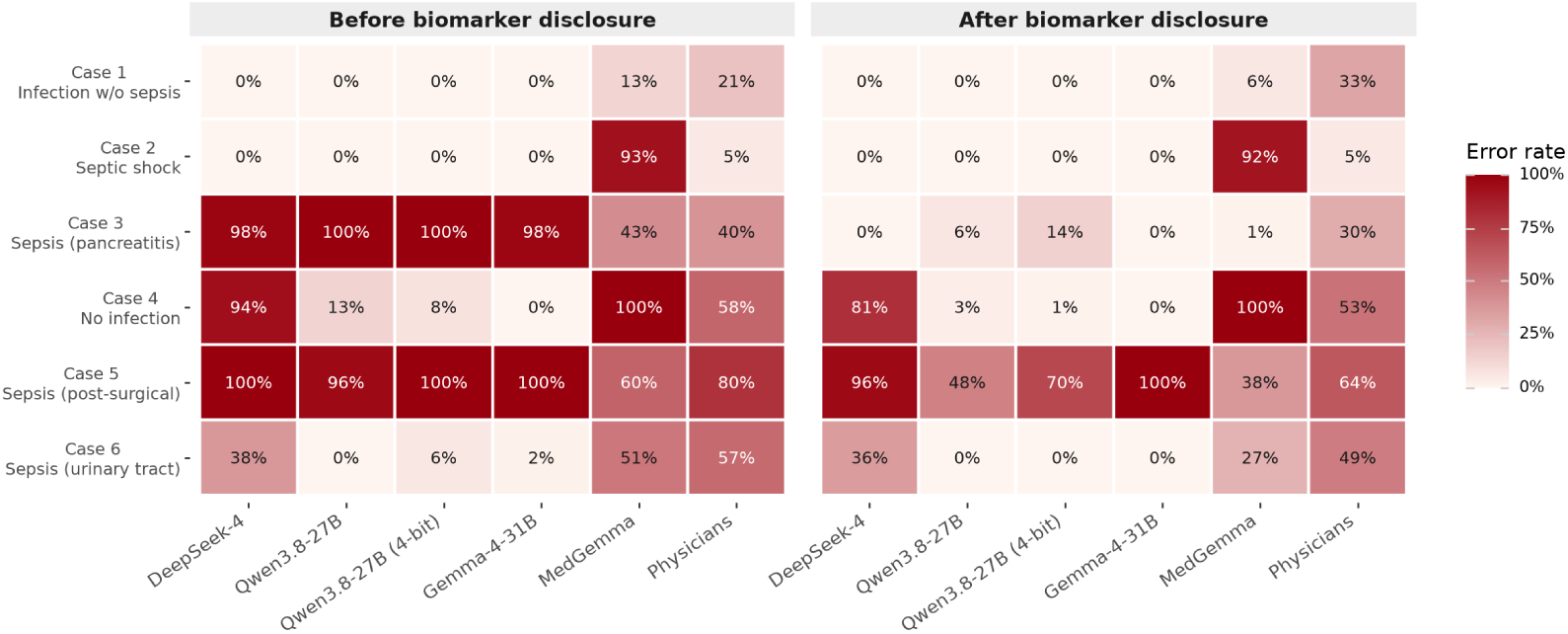
Diagnostic error rate by clinical vignette. Each cell shows the diagnostic error rate for a given model on a given clinical vignette; darker shading indicates a higher error rate. For the language models, the error rate is pooled across all experimental configurations of that vignette (patient sex, age, language, reasoning mode and sampling temperature), with the three repetitions of each configuration collapsed by majority vote. The rightmost column (Physicians) shows the observed per-vignette error rate of the human comparator (N = 353) from the companion survey, included as a baseline.

Two further analyses characterise the models’ behaviour. First, each configuration was run three times to assess diagnostic stability under stochastic decoding (Appendix B): Gemma-4-31B was the most reproducible model, maintaining near-perfect agreement across the three repetitions even at sampling temperatures of 0.3 and 0.7, where the other models showed a marked decline. Second, we examined whether accuracy depended on the diagnostic framework each model invoked to justify its decision, grouping configurations by the framework cited at stage 1 — Sepsis-3 (SOFA/qSOFA), Sepsis-2 (SIRS), a mixed Sepsis-2/3 rationale, or expert clinical judgment without an explicit framework (Appendix B). This relationship was strongly model-dependent, with no single framework uniformly associated with higher accuracy across models.

### 3.2. Bias Analysis

Figure 3 summarises each model’s sensitivity to variations in patient sex, age, language and reasoning mode — factors that were varied while holding the clinical information constant. Sensitivity is quantified as the diagnosis-change rate: the proportion of configurations identical in every respect except the factor of interest in which the voted diagnosis changed when only that factor was varied. Sampling temperature, which should likewise not affect the diagnosis, serves as a per-model noise reference.

**Figure 3:**
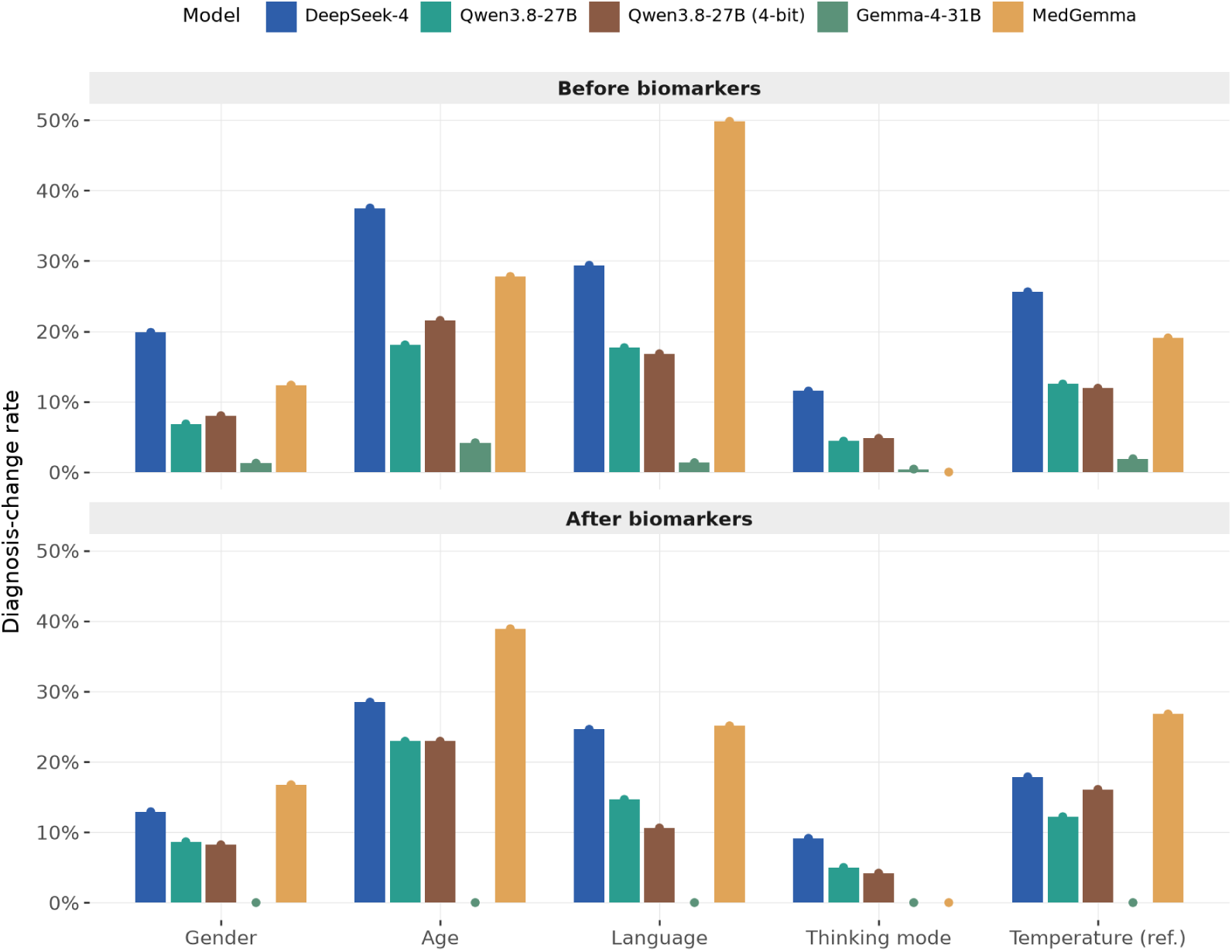
Sensitivity of each model to attributes that should not change the diagnosis, before and after biomarker disclosure. For each attribute, configurations identical in every respect except that attribute were grouped, and the diagnosis-change rate was computed as the proportion of groups in which the majority-voted diagnosis (over the three repetitions) changed when only that attribute was varied — for example, a gender bar of 13% means that in 13% of otherwise-identical male–female configuration pairs the model’s voted diagnosis differed solely because the patient’s gender was changed. MedGemma has no reasoning-mode toggle, so its “Thinking mode” bar is absent.

Gender and reasoning mode remained at or below the noise level in every model, whereas age and language were the factors that most often exceeded it. Gemma-4-31B was again the most robust model, with all bars close to 0%, while DeepSeek-4 and MedGemma were the most unstable. Qwen3.8-27B and its quantised variant showed moderate and nearly identical profiles, with age and language slightly above their noise floor (12–13%), indicating that quantisation did not substantially alter their sensitivity.

High instability does not by itself imply bias. Figure 4 contrasts, for each model and attribute, the diagnosis-change rate with the accuracy spread across that attribute’s levels. Sex produced flip rates (up to 17%) but near-zero accuracy spread, indicating no directional sex effect. Age generated the largest instability (flip up to 39%) yet a spread of 6–11%, so most of this variability was non-directional noise rather than a systematic age gap. Language, in contrast, lay closest to the diagonal (most clearly for DeepSeek-4 and the Qwen3.8-27B) showing that language-induced instability did translate into a real accuracy gap. Gemma-4-31B clustered at the origin for all three attributes, confirming it as the most robust model, whereas DeepSeek-4 and MedGemma were the noisiest.

**Figure 4:**
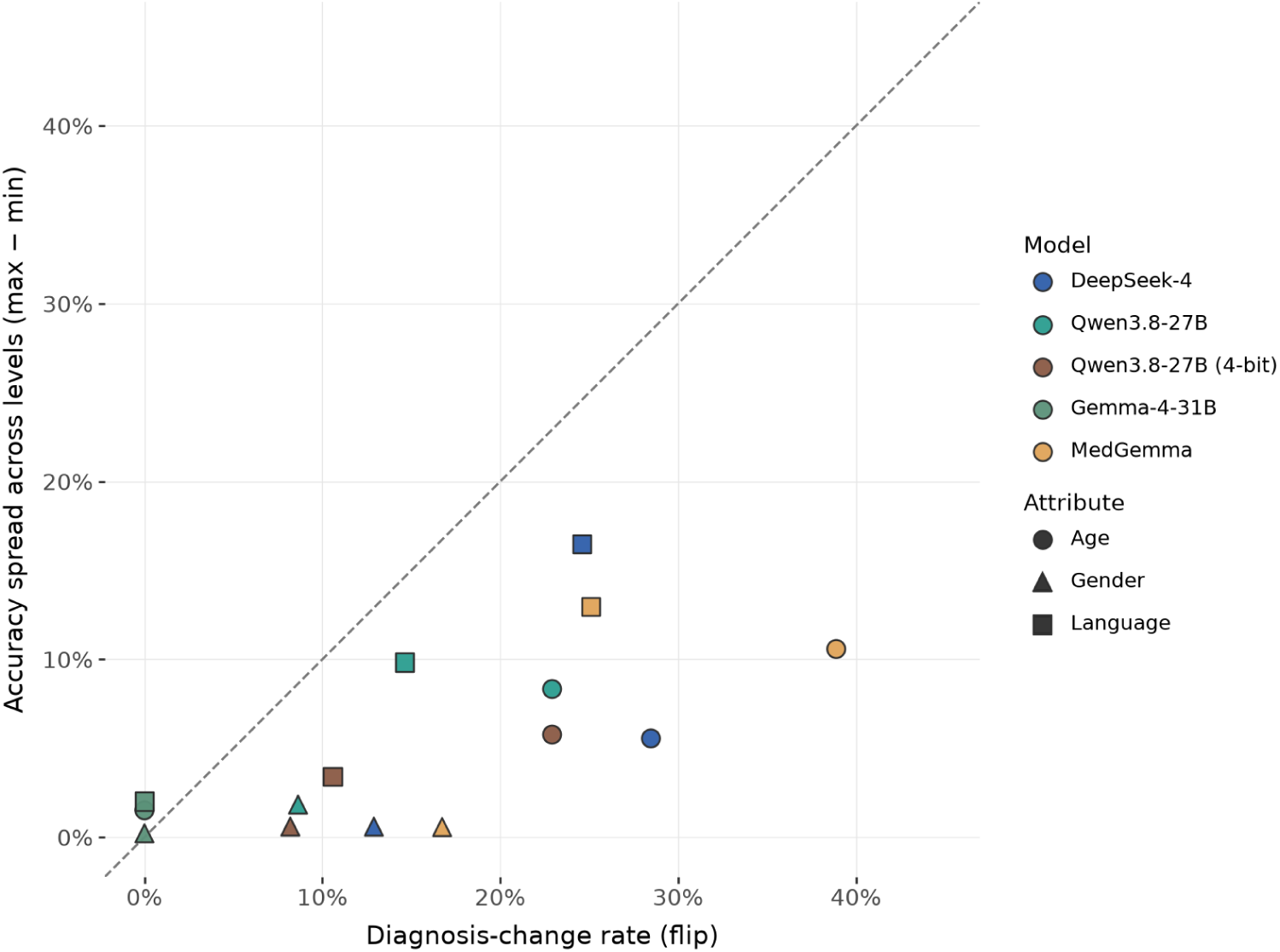
Instability versus directional bias for each model. Each point represents one model-attribute combination (circle = Age, triangle = Sex, square = Language, colour = model). The x-axis is the diagnosis-change rate (flip): the proportion of otherwise-identical configuration pairs in which the majority-voted diagnosis changes when only that attribute varies, i.e., how unstable the model is to the attribute. The y-axis is the accuracy spread across the attribute’s levels (maximum - minimum accuracy), i.e., how much that instability translates into a systematic performance gap.

Finally, we assessed whether stated confidence tracked actual accuracy, which for a well-calibrated model should increase monotonically from low to very-high confidence. As shown in Figure 5, the two Qwen models approached this ideal most closely: Qwen3.8-27B rose from 75% accuracy at moderate confidence to 98% at high and 100% at very high, and its quantised variant from 61% to 100%, so their stated confidence tracked their performance. DeepSeek-4 was also monotonic. Gemma-4-31B, by contrast, was non-monotonic and poorly ordered, and MedGemma was the most miscalibrated.

**Figure 5:**
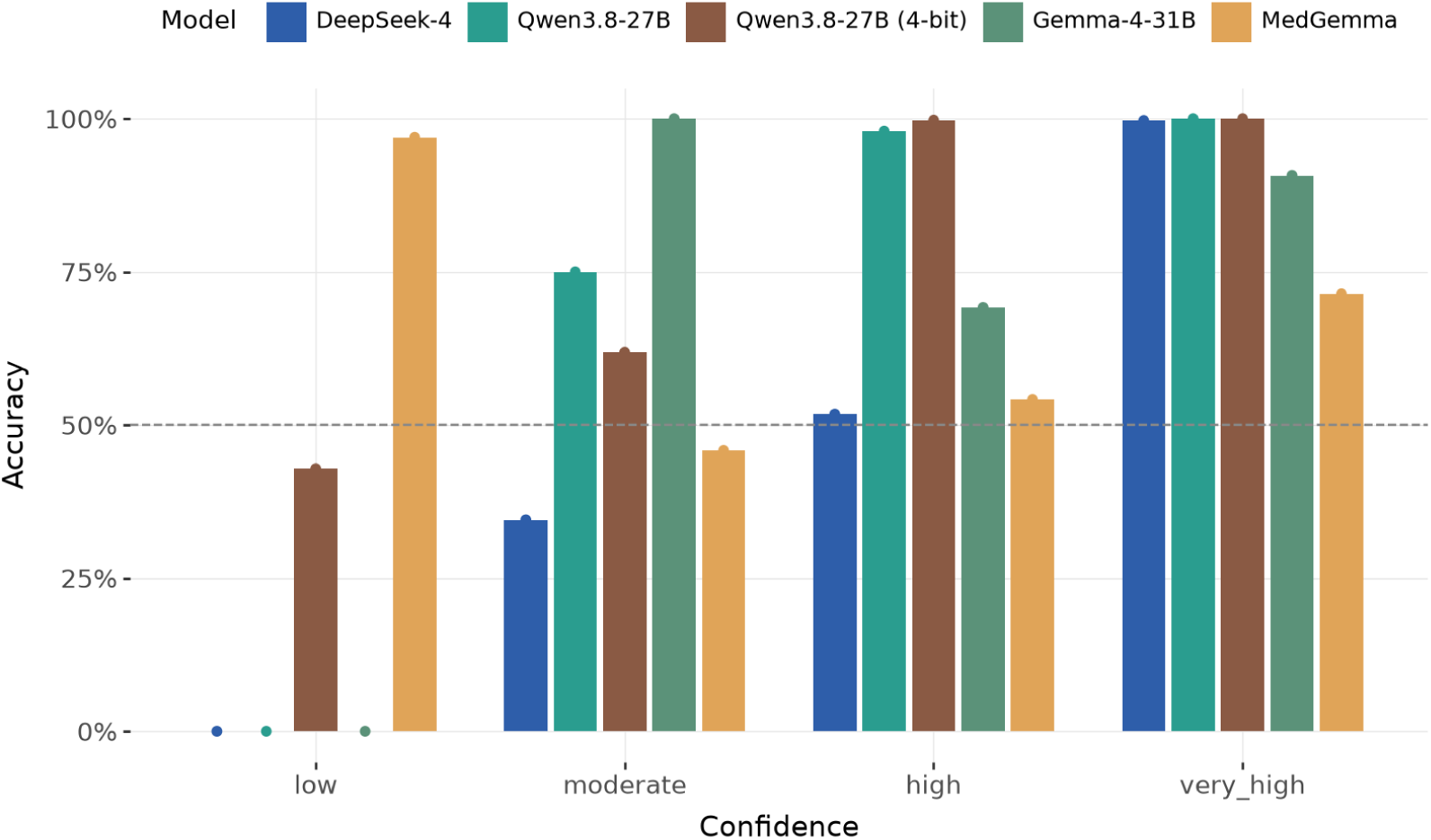
Calibration of each model: diagnostic accuracy within each self-reported confidence level after biomarker disclosure. For every model, final diagnoses were grouped by the confidence the model stated alongside them, and the bars show the proportion of correct diagnoses (against the Sepsis-3 reference standard) within each confidence level, coloured by model. A well-calibrated model should show accuracy increasing monotonically from left (low confidence) to right (very high confidence), so that higher stated confidence corresponds to higher actual accuracy.

## 4. Discussion

In this evaluation, five open-weight LLMs were assessed on six standardised sepsis vignettes drawn from unpublished, de-identified clinical records, which were therefore unlikely to have formed part of the models’ training corpora. Before biomarkers, our models were broadly comparable to physicians (40–67% vs 57%), but after disclosure the strongest models surpassed the physician baseline (83–91% vs 61%). This is consistent with reports that frontier models can match or exceed clinicians on structured diagnostic tasks [26], while also echoing the more nuanced view that LLMs excel at confirmatory reasoning but struggle with early or ambiguous presentations [27].

Our case-level analysis makes this mechanism explicit: errors concentrated in the sepsis cases that require laboratory confirmation (acute pancreatitis, post-surgical sepsis), and biomarker disclosure produced a case-specific rescue — the error rate for the pancreatitis case collapsed from 98%-100% to 0–14%. The persistence of post-surgical sepsis as the hardest vignette for both models and physicians (80% → 64% error) suggests intrinsic case difficulty rather than a model-specific failure. Whereas most prior diagnostic evaluations rely on proprietary systems [28], our study focuses exclusively on open-weight models, which is increasingly relevant given privacy and deployment constraints in clinical settings. Our finding that the Qwen family were among the strongest models aligns with recent systematic evaluations of open-weight LLMs for clinical reasoning [29]. Importantly, quantisation of Qwen3.8-27B incurred only a small accuracy cost (91% → 86%) and did not substantially alter its bias or sensitivity profile. Across the bias analysis, patient sex and reasoning mode behaved as noise, age produced instability without a systematic accuracy gap, and only language emerged as a directional bias. This pattern complements the growing body of fairness benchmarks for medical LLMs, which consistently detect residual demographic disparities even in the least biased systems [30, 31, 32]. This is particularly relevant for the multilingual deployment scenario, and argues for language-specific validation before clinical use. A central message of our study is that accuracy, robustness and calibration are distinct dimensions that do not covary: Gemma-4-31B was the most reproducible and least biased model yet not the most accurate, whereas the most accurate Qwen models were noisier. This dissociation echoes recent work cautioning that benchmark accuracy can mask latent safety and stability problems [33], and that reproducibility across repeated runs is itself a measurable property of clinical LLMs. Notably, the only domain-specific model, MedGemma, was the weakest and least calibrated, with an anomalous error profile that did not follow the clinical difficulty gradient of the other models. This should be interpreted with caution: MedGemma is built on an architecturally older backbone than the other models evaluated, so its underperformance more likely reflects its model generation than a failure of medical specialisation per se. This caveat is consistent with evidence that newer general-purpose models can outperform earlier domain-tuned systems, and it motivates re-evaluating medical fine-tuning on top of current-generation backbones.

## 5. Limitations

Our study has several limitations. It is an in-silico evaluation on six standardised vignettes rather than prospective, real-world electronic health record data, so absolute accuracies may not transfer to clinical practice, and aggregate results are sensitive to individual cases. The physician comparator is drawn from a companion survey in which each vignette was assessed once in its original form, so the comparison with the models, pooled across varied configurations, is not fully like-for-like. Models were queried zero-shot with a single prompt. The bias analysis covered only sex, age and two languages. The per-model configuration counts differ (MedGemma lacks a reasoning-mode toggle and uses an older backbone), quantisation was assessed for a single model family.

## 6. Conclusion

In this evaluation on six standardised sepsis vignettes, LLMs matched practising physicians before biomarker disclosure and surpassed them afterwards, with the strongest models (Qwen3.8-27B and Gemma-4-31B) reaching 83–91% accuracy versus 61% for physicians. Robustness and calibration varied substantially: Gemma-4-31B was the most stable and least biased model, the Qwen family was the best calibrated, and the quantisation preserved most of the diagnostic performance at a negligible cost, supporting the feasibility of resource-efficient deployment. Across the bias analysis, sex and age behaved as noise and only language emerged as a directional bias, underscoring the need to validate these systems across languages before clinical use. Although MedGemma is the only medically specialised model in our panel, it ranked lowest in both accuracy and calibration. We interpret this cautiously, since its backbone predates that of the other models. Overall, general-purpose LLMs show promise as decision support system for sepsis, yet their readiness for clinical use depends as much on robustness, calibration and language fairness as on raw accuracy.

## Data Availability

All data produced in the present study are available upon reasonable request to the authors

## Appendix A. Clinical Cases

**Table A.1:**
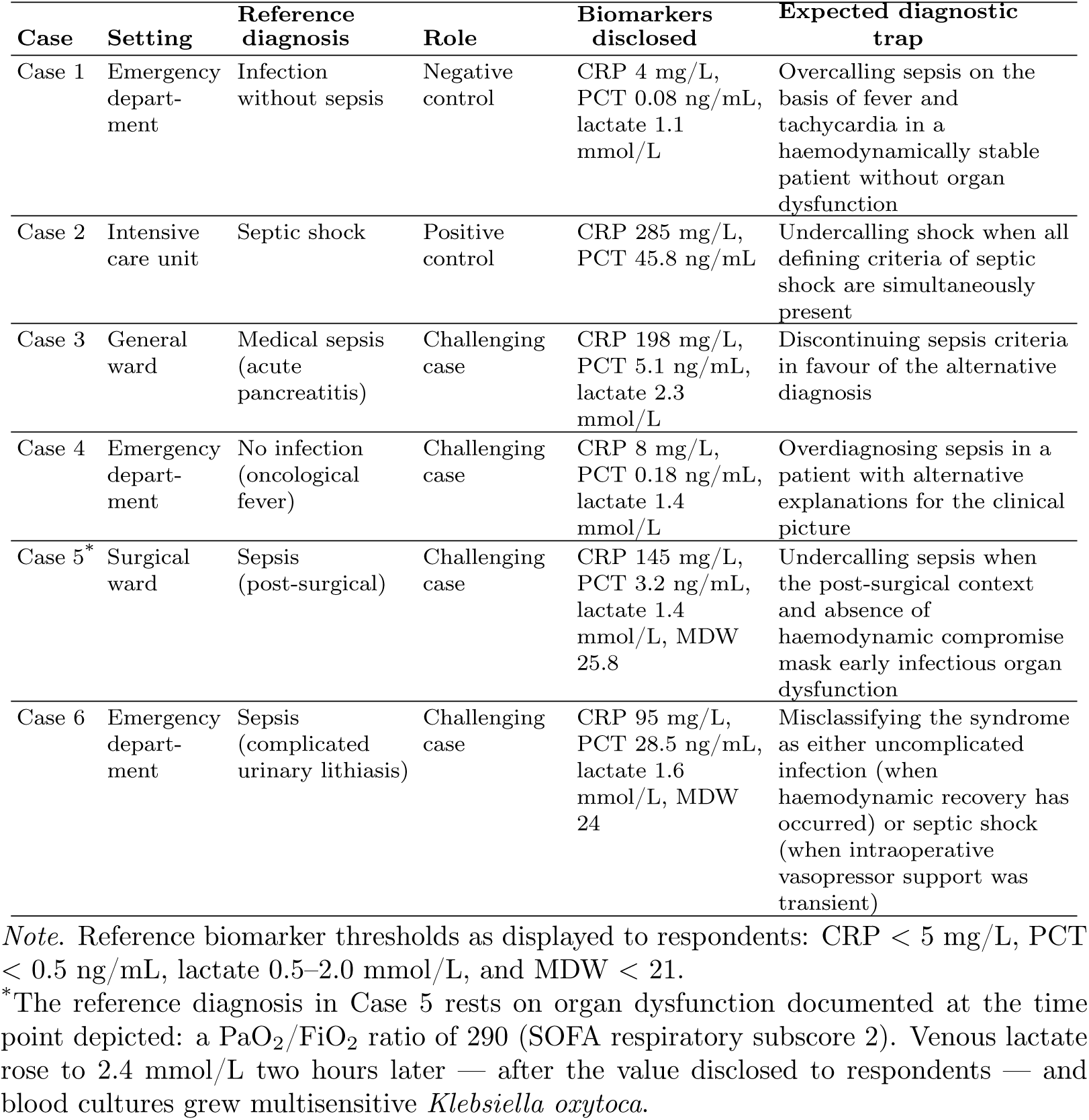
Description of the six clinical vignettes used in the survey.

## Appendix B. Reproducibility and Diagnostic-Framework Effects

**Figure B.6:**
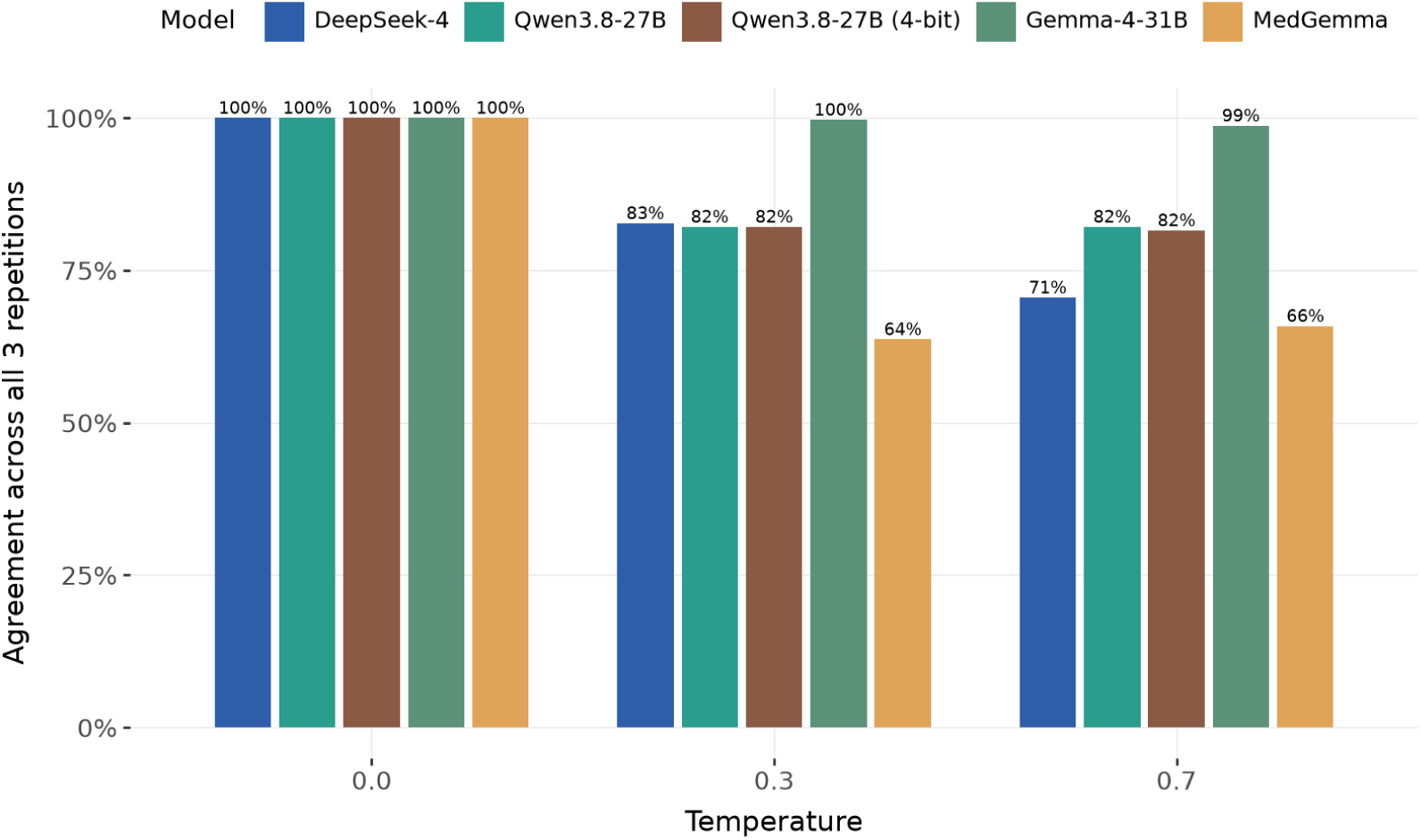
Reproducibility. Each configuration was run three times, bars show the proportion with identical final diagnoses across all three repetitions, per model and temperature (0.0, 0.3, 0.7). Determinism at temperature 0 should yield 100%; the drop at higher temperatures reflects sampling stochasticity.

**Figure B.7:**
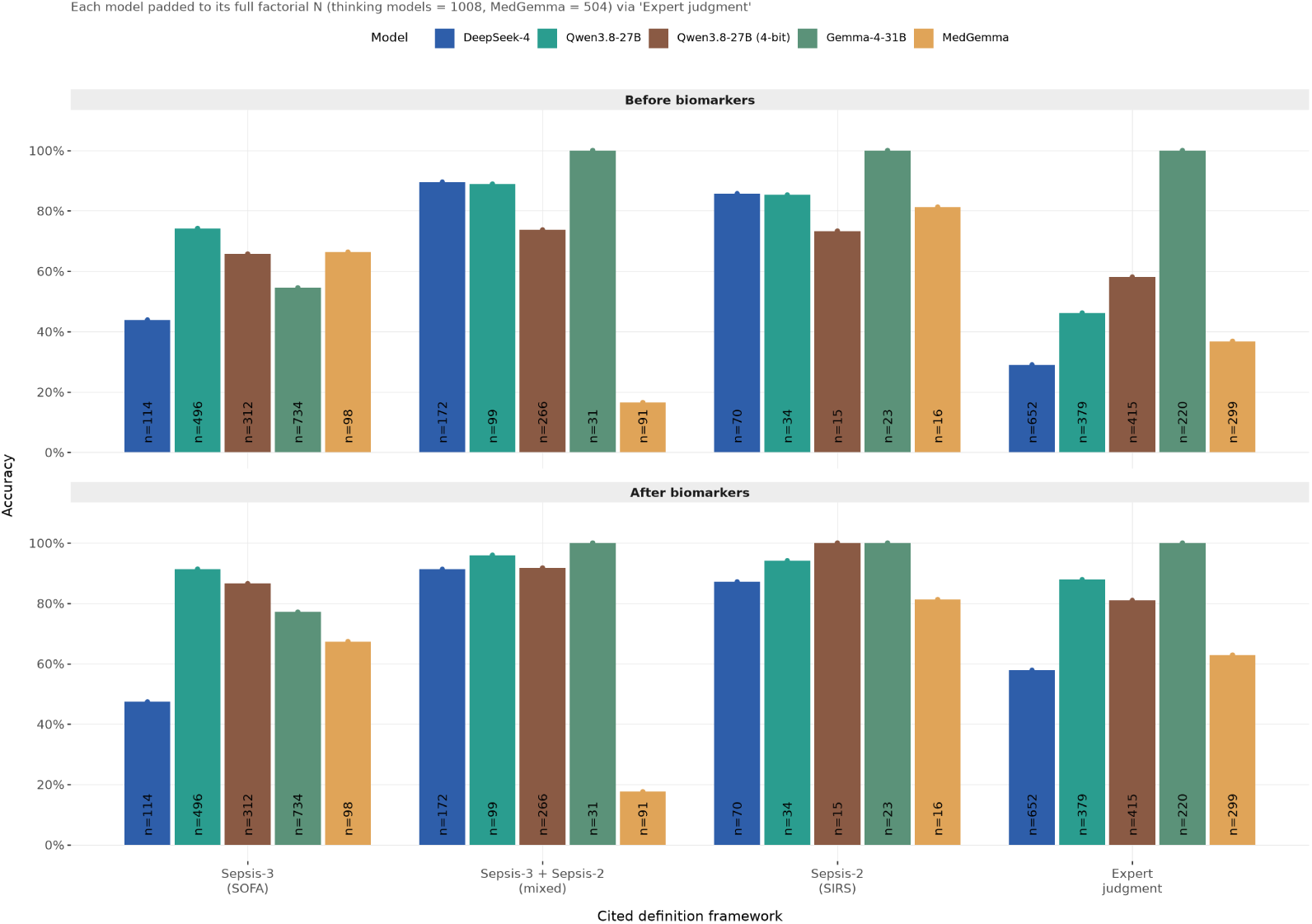
Diagnostic accuracy by the sepsis-definition framework cited by each model. Each model’s configurations were grouped by the diagnostic framework it reported at stage 1, assigned once per configuration by majority vote of the three repetitions: Sepsis-3 (SOFA/qSOFA), a mixed Sepsis-3 + Sepsis-2 rationale, Sepsis-2 (SIRS), or expert/clinical judgment with no formal framework.

Accuracy varied substantially with the framework a model cited, and the pattern was model-specific. DeepSeek-4 was least accurate precisely in its two most frequent rationales — Sepsis-3/SOFA (44% before, 47% after biomarkers) and expert judgment (29% → 58%; n = 652) — while performing well whenever it invoked a Sepsis-2 (SIRS) or mixed rationale (86–91%). Qwen3.8-27B and its quantised variant showed the clearest benefit of biomarker disclosure within the SOFA bucket. Gemma-4-31B was close to 100% across the mixed, SIRS and expert-judgment buckets but comparatively weaker in its dominant SOFA bucket (54% → 77%; n = 734). Overall, biomarker disclosure improved accuracy within almost every framework, with the largest gains concentrated in the SOFA and expert-judgment categories.

